# Digital game-based interventions for cognitive training in healthy adults and adults with cognitive impairment: Protocol for a two-part systematic review and meta-analysis

**DOI:** 10.1101/2022.12.13.22283401

**Authors:** Shi-Bei Tan, Joshua Tan, Marlena N. Raczkowska, Joshann Chean Wen Lee, Bina Rai, Alexandria Remus, Dean Ho

**Author notes:** Contact details: Both corresponding authors can be reached at The N.1 Institute for Health (N.1), National University of Singapore, Singapore, 117456, Singapore; (Alexandria Remus); (Dean Ho). These authors contributed equally (Alexandria Remus, Dean Ho).

## Abstract

**Introduction:** Digital game-based training interventions are scalable solutions that may improve cognitive function for many populations with varying abilities. This two-part review aims to synthesise the effectiveness and key features of digital game-based interventions for cognitive training in: 1) healthy adults across the life span; and 2) adults with cognitive impairment, to update current knowledge and impact the development of future interventions for different adult subpopulations.

**Methods and analysis:** This systematic review protocol follows the Preferred Reporting Items for Systematic Reviews and Meta-Analyses Protocols guidelines. A systematic search was performed in PubMed, Embase.com, CINAHL, Cochrane Library, Web of Science, PsycINFO and IEEE Explore on July 31^st^, 2022 for relevant literature published in the previous five years. Experimental, observational, exploratory, correlational, qualitative and mixed-methods studies will be eligible if they report at least one cognitive function outcome and include a digital game-based intervention intended to improve cognitive function. Reviews will be excluded but retained to search their reference lists for other relevant studies. All screening will be done by at least two independent reviewers. The appropriate Joanna Briggs Institute Critical Appraisal Tool, according to the study design, will be applied to perform the risk of bias assessment. Outcomes related to cognitive function and digital game-based intervention features will be extracted. Results will be categorised by adult life span stages in the healthy adult population for Part 1 and by neurological disorder in Part 2. Extracted data will be analysed quantitatively and qualitatively, according to study type. If a group of sufficiently comparable studies is identified, we will perform a meta-analysis applying the random effects model with consideration of the I^2^ statistic.

**Ethics and dissemination:** Ethics approval is not applicable for this study since no original data will be collected. The results will be disseminated through peer-reviewed publications and conference presentations.

**PROSPERO registration number:** CRD42022351265

**Strengths and limitations of this study:**

- We will adhere to the rigorous methodology in accordance with the most recent Preferred Reporting Items for Systematic Reviews and Meta-Analyses guidelines.
- The search strategy was developed in consultation with an experienced research librarian and customised to eight relevant databases.
- The focus on cognitive outcomes and digital game-based intervention features will allow for in-depth analysis and insights for developing future interventions for specific populations.
- This systematic review may be limited in power for generalisation if a limited number of studies are reported either for each adult lifespan stage or for each neurological disorder of interest.
- The English language restriction may exclude relevant studies reported in a non-English medium.

## Introduction

As we age, cognitive function improves early in life, but our abilities begin to decline as we progress through the later stages of the adult life span. Whilst cognitive impairments associated with normal ageing may manifest in adults, these impairments are generally not so severe that they negatively impact daily functioning and Quality of Life (QoL)^4^. However, various abnormal aging and neurological disorders prevalent throughout the different phases of the adult span are known to result in cognitive impairments, which in turn may reduce self-reported QoL in some individuals^2,3^. In fact, reduced cognitive function is reported to be associated with lower self-reported QoL in older adults with subjective cognitive impairment^1^, mild cognitive impairment (MCI)^2^, dementia^2,3^, stroke^5^ and high-grade glioma^6^. As the global population ages and life expectancy increases, the incidence of such neurological disorders with associated cognitive impairment will also rise. For example, the already high incidence of dementia is well accepted with new cases of Alzheimer’s disease in the United States forecasted to double by 2050^7^ and improving survival rates post-stroke expected to lead to an increase in vascular and post-stroke dementias^8,9^. Further, it is expected that MCI could double the mortality risk^10^. Thus, with the aging global population on the rise, identifying effective strategies for improving cognitive function in both healthy adults and those with cognitive impairment is increasingly important.

In healthy adult populations, there is evidence that cognitive training can improve cognitive functions, suggesting that the aging brain is still amenable to neuronal and cognitive plasticity^11^. It is also thought that cognitive training may delay the onset and progression of the cognitive impairment associated with some neurological disorders, which is projected to significantly reduce the global burden of the disease^12^. With mobile technologies being widely available, affordable and popular^13^, there is great potential to harness digital game-based training for cognitive health. In fact, numerous studies have shown potential benefits of digital games on health, especially for mental health^14,15^, and there has been an enormous increase in interactive software programmes created with claims of their ability in improving fundamental aspects of cognition over the last decade^16^. While some studies on digital game-based cognitive training highlighted controversial results^17,18^, others have demonstrated promising improvements across several cognitive outcomes, specifically multiple domain, processing speed and reaction time, memory, task-switching/multitasking, mental spatial rotation, top-down attention and spatial cognition^13,19,20^, which highlights the potential in digital game-based cognitive training.

While multiple systematic reviews and meta-analyses investigating the effects of digital game-based interventions on cognitive functioning for specific age groups^21,22^ or conditions^18^ have been published, there is a lack of similar evidence syntheses of such investigations in healthy individuals by stages of the adult life span and in adults by neurological disorders with associated cognitive impairment. Further, these published reviews with specific groups are siloed, making it challenging for future game-based intervention developers to be well-informed. For example, one review published in 2018^13^ synthesised the evidence of video game training on cognitive and emotional skills in a healthy adult population from 18 to 59 years old published between 2013 – 2018, but did not explore effects by age groups across the lifespan. Moreover, the COVID-19 pandemic, starting in late 2019, gave rise to a significant increase in digital health technologies^23^, therefore a comprehensive review including recent digital game-based interventions from the last few years is warranted.

With this in mind, we aim to perform a systematic review of the recent literature of digital game-based cognitive training interventions from the last five years. Specifically, our research questions are: 1) What is the effectiveness of digital game-based interventions for improving cognitive function?; and 2) What are the key features of the digital game-based cognitive interventions?. We will explore these research questions in two broad population groups: 1) healthy adults across the adult life span; and 2) adults with neurological disorders with associated cognitive impairment. We aim to report the results in a two-part review – one for each of the populations mentioned – and perform a meta-analysis, if sufficiently comparable studies are identified.

## Methods

This systematic review will be conducted and reported following the Preferred Reporting Items for Systematic Reviews and Meta-Analyses (PRISMA) guidelines^24^. This protocol adheres to the PRISMA-P (PRISMA Protocols) reporting guidelines^25^.

## Eligibility criteria

### Type of studies

Experimental, observational, (exploratory, correlational, qualitative (where a cognitive training game is being developed and/or assessed for acceptability, feasibility, usability etc.) and mixed-methods studies will be eligible as long as they report one of the five cognitive domains^26^ (namely executive function, perceptual-motor function, language, learning and memory, and complex attention) and include at least one digital game-based intervention intended for cognitive training. Reviews will be excluded but retained in order to search their reference lists for other relevant studies. Randomised control trials will be considered for meta-analyses and all other study types will be synthesised narratively. We will include studies from all settings (e.g., home, community, hospital ward, long-term care facilities, etc.).

### Types of participants

For Part 1 of the review, participants will include healthy adults (18+) across all stages of the adult life span will be studied. For Part 2 of the review, participants will include adults with prevalent neurological disorders with associated cognitive impairment, specifically: MCI, dementia, Alzheimer’s disease, stroke, and brain tumours, will be studied.

### Type of interventions

Digital game-based interventions intended for cognitive training, defined as any computerised tasks for the specific purpose of improving at least one cognitive function: executive function, perceptual-motor function, language, learning and memory, and complex attention.

### Type of control

For relevant study designs, the comparison intervention can include passive or active controls digital game-based interventions, placebo intervention, no intervention or standard of care.

### Type of outcomes

Our primary outcome measures will be cognitive function and digital game-based intervention features to address our research questions. Our secondary outcome measures will include study design/implementation features and any potential game-based digital biomarkers for cognitive function. Specifically, digital biomarkers will be defined as objective, quantifiable physiological and behavioural data collected from the digital game-based intervention that may offer the ability to detect changes in cognitive function^27^.

## Information sources

A systematic search was performed in PubMed, Embase.com, CINAHL, Cochrane Library, Web of Science, PsycINFO and IEEE Explore on July 31^st^, 2022 to search for literature published from January 2017 until July 2022. A customised search strategy following the PICO (Population, Intervention, Control, and Outcomes) method was conducted for each database in consult with a research librarian and tailored according to each database requirements. The searches were limited to publications in the English language. Where applicable, type of publication was limited to academic journals, articles in press, conference papers, but not abstracts or trial registrations. The reference lists of studies meeting the inclusion criteria will be searched to identify additional relevant studies. After full text screening, eligible articles will be separated by Part 1 and Part 2 populations for analyses.

## Search strategy

The search for PubMed is provided as an example and is available in online supplemental Appendix 2.

The search terms used were:

1. Digital games *Types of games*: Digital, online, video, computer games, interactive, serious, mobile, tablet, app-based, web-based, handheld, console, commercial, multiplayer, single-player, video, mobile, simulation, cognitive, training *Gaming genre*: platforming, shooters, strategy games, racing games, real-time strategy, simulation and sports, survival, horror, puzzles, party, etc. Game device: digital device, smartphone, tablet, simulator, Kinect, computer, virtual reality, handheld device, console, controllers, keyboard, augmented reality
2. Cognitive training and functioning: cognitive training, cognition, cognitive task, memory, learning, executive function, language, perceptual-motor function, complex attention, attention, sustained attention, divided attention, selective attention, processing speed
3. Adult: adults, 18+, seniors, elderly, middle-aged, early middle-aged, late middle-aged, young adults
4. (1) AND (2) AND (3)

The search was built by consultation an experienced research librarian at the XX and incorporated controlled vocabulary terms for each specific database searched.

### Data records and management

Initial searches were performed by one reviewer. All duplicate removal and screening will be completed using Covidence (a web-based systematic review software available at www.covidence.org). To date, at least two reviewers have applied eligibility criteria to titles and abstracts. At least two reviewers will next apply eligibility criteria to the full-text articles Any discrepancies will be addressed by a third independent reviewer. Reasons for exclusion of studies will be recorded during the full-text screening phase. Two reviewers will independently extract study data, with findings compared and agreed. A bespoke data extraction spreadsheet (Excel) will be adapted and modified from Pallavicini et al^13^. Data will be coded into three broad categories: (1) cognitive function outcomes; (2) digital game-based intervention outcomes; and (3) study outcomes. Cognitive function outcomes include but are not limited to domain; memory; working memory; episodic memory; near transfer; far transfer; general intelligence; processing speed; reaction timing; task-switching/multi-tasking; mental spatial rotation; inhibition control; attention; executive function; visuospatial function; language; speed-attention; numeric reasoning; potential digital biomarkers; global cognition; quality of life. Digital game-based intervention outcomes include but are not limited to game category; game genre; platform of delivery; number of players; primary purpose of the game; gamification features; game description; duration of intervention; frequency of intervention; prescription of game. Study outcomes include but are not limited to: abstracts, aim of study; study design; population; diagnosis and inclusion/exclusion criteria for patients (for Part 2); course and duration of conditions/symptoms (for Part 2); other medical diagnoses; number of patients: screened, eligible, ineligible, enrolled, randomised to intervention, randomised to control, excluded post randomisation, withdrawn, lost to follow up, included in analysis and each outcome measures; age; gender; geographic location (city/state/country); ethnicity; measures used for cognitive assessment; setting of training; adverse events. If data to be extracted are missing, incomplete or unclear, inquiries will be sent to the authors.

### Effect measures

We will report the treatment effects between the intervention and control group as the mean difference for continuous data and risk ratio for dichotomous data with accompanying 95% confidence intervals (CI).

For continuous data on target outcomes, a generic inverse variance random effects model will be used to pool the mean difference (MD) with 95% CI. It is anticipated that the units of the outcome measures used across studies may not be consistent and therefore it is likely that we will report the effects as standardised mean differences (SMD) rather than MD. For dichotomous data, a random effects method will be used to pool the summary risk ratio (RR) with 95% CI. An overall effect size with 0.2-0.5 will be regarded as small, 0.5-0.8 as moderate and more than 0.8 as large^28^. If means but not standard deviations have been reported, we will attempt to calculate the standard deviation from the information reported.

### Risk of bias

Two reviewers will assess the risk of bias, independently. Due to the heterogeneity of possible included studies, we cannot state beforehand if one specific tool to assess the risk of bias is sufficient. Therefore, we will use the appropriate Joanna Briggs Institute Critical Appraisal Tool (JBICAT) at the study level according to the study design, retrieved from https://jbi.global/critical-appraisal-tools. If disagreements between reviewers arise, they will be solved either through discussion or with the help of a third reviewer. Answering all questions of JBICAT will lead to a classification for each study, following the system of classification of low risk of bias (all criteria are met), moderate risk of bias (two criteria are not met or remain unclear) or high risk of bias (three or more criteria are not met or remain unclear).

### Data synthesis

For Part 1 of the review, individual studies will be combined and grouped by the different stages across the healthy adult life span, which is anticipated to be young adults, middle-aged adults, and elderly. For Part 2 of the review, individual studies will be combined and grouped by the neurological disorder of interest, specifically MCI, dementia, Alzheimer’s, stroke and brain tumours. All statistical tests and overall effect sizes will be conducted and estimated using Review Manager 5 (RevMan 5), Version 5.4 (The Cochrane Collaboration, 2020).

First, a narrative synthesis and descriptive statistics will be provided in the text and tables to summarise the study characteristics and results. If further quantitative synthesis is appropriate, we will perform a meta-analysis on groups of studies with sufficiently comparable intervention and cognitive outcome, if identified, by applying the intention-to-treat principle. We will assess between study heterogeneity using the I^2^ statistic which describes the percentage of variability in effect estimates due to heterogeneity rather than chance. Thresholds for I^2^ will be >30% for moderate heterogeneity, >50% for substantial heterogeneity and >75% for considerable heterogeneity. Sensitivity analysis will be conducted according to overall study quality which will be split into three categories of low risk of bias, some concerns, and high risk of bias, by comparing random and fixed-effect models and by excluding possible outlying studies, e.g., if the visual inspection of the forest plot shows poorly overlapping CIs. The possibility of publication bias will be explored by constructing funnel plots and by conducting the Egger’s test^29^ for analyses that contain more than 10 studies.

### Confidence in cumulative evidence

The quality of the effect estimates for each reported outcome will be assessed using the Grading of Recommendations, Assessment, Development and Evaluation (GRADE) approach^30-32^ by two reviews. Possible disagreements will be assessed by a third review.

### Ethics and dissemination

Ethics approval is not applicable for this study since no original data will be collected. The results will be disseminated through peer-reviewed publication and conference presentations.

### Patient and public involvement

Patients or the public were not involved in the design, or conduct, or reporting, or dissemination plans of our research.

## Discussion

While digital game-based cognitive training interventions may be beneficial in improving outcomes in the adult population, there is a lack of investigation of such effects by age group and by neurological disorders with associated cognitive impairment. Greater understanding of the effectiveness and features of these interventions based on different subpopulations will be useful to inform developers of such interventions for specific subpopulations.

This systematic review and meta-analysis will update the existing knowledge on the effectiveness and key features of digital game-based interventions for cognitive training, with comprehensive analysis in terms of healthy adults across the phases of the adult life span and adults with cognitive impairment. The results may impact future development of digital game-based interventions for cognitive training for different subpopulations of adults but may be limited by the possibly low number and quality of the research studies available.

## Data Availability

All data produced in the present work are contained in the manuscript

## Ethics statements

Ethics approval is not applicable for this study since no original data will be collected.

## Patient consent for publication

Not applicable.

## Authors’ contributions

AR, MNR, S-BT, and JT designed the protocol. AR and S-BT wrote the first draft of the protocol. AR, MNR, S-BT, and JT designed the search and S-BT and JT performed the searches. All authors provided critical appraisal regarding the design of the systematic review and revised the manuscript. All authors approved the final version of the protocol.

## Funding Statement

DH gratefully acknowledges funding from the Singapore Cancer Society [grant number SCS-GRA-2019-00063], the National Research Foundation Singapore under its AI Singapore Programme [Award Number: AISG-GC-2019-002], and the Singapore Ministry of Health’s National Medical Research Council under its Open Fund-Large Collaborative Grant (“OF-LCG”) [grant number MOH-OFLCG18May-0028], Institute for Digital Medicine (WisDM) Translational Research Programme [grant number R-719-000-037-733] at the Yong Loo Lin School of Medicine, National University of Singapore, Ministry of Education Tier 1 FRC Grant [grant number R-397-000-333-114] and the Next-Generation Brain-Computer-Brain Platform – A Holistic Solution for the Restoration & Enhancement of Brain Functions (NOURISH) project from the RIE2020 ADVANCED MANUFACTURING AND ENGINEERING (AME) PROGRAMMATIC FUND [grant number A20G8b0102 / A-0002199-02-00]. Note that none of the funders stated above have influenced or provided inputs on the design or scope of this systematic review.

## Competing Interest Statement

DH and SB-T are co-inventors of previously filed pending patents on artificial intelligence-based therapy development. DH is a shareholder of KYAN Therapeutics, which has licensed intellectual property pertaining to AI-based oncology drug development and personalised medicine.

[Link to online supplemental Appendix 1 and 2]

## Notes

### Funding Statement

DH gratefully acknowledges funding from the Singapore Cancer Society, the National Research Foundation Singapore under its AI Singapore Programme, and the Singapore Ministry of Healths National Medical Research Council under its Open Fund- Large Collaborative Grant,  Institute for Digital Medicine (WisDM) Translational Research Programme at the Yong Loo Lin School of Medicine, National University of Singapore, Ministry of Education Tier 1 FRC Grant and the Next Generation Brain Computer Brain Platform A Holistic Solution for the Restoration & Enhancement of Brain Functions (NOURISH) project from the RIE2020 ADVANCED MANUFACTURING AND ENGINEERING (AME) PROGRAMMATIC FUND. Note that none of the funders stated above have influenced or provided inputs on the design or scope of this systematic review.

