## Appendix 2 for "Digital game-based interventions for cognitive training in healthy adults and adults with cognitive impairment: Protocol for a two-part systematic review and meta-analysis"

| Search number | Query |
| --- | --- |
| 1 | "Adult"[Mesh] |
| 2 | Adult* OR senior* OR elder* OR old OR (middle aged OR middle-aged) |
| 3 | "Video Games"[Mesh] |
| 4 | ("Digital Technology"[Mesh] OR "Computers"[Mesh] OR "Computers, Handheld"[Mesh] OR "Simulation Training"[Mesh] OR "Sports"[Mesh]) OR ("Cell Phone"[Mesh] OR "Computers, Handheld"[Mesh] OR "Smartphone"[Mesh] OR "Augmented Reality"[Mesh] OR "Virtual Reality"[Mesh]) |
| 5 | ((Digital OR online OR training OR video OR interactive OR serious OR commercial OR multiplayer OR single-player OR simulation OR platform* OR shoot* OR strategy OR racing OR sports OR survival OR horror OR puzzle OR party) OR (mobile OR tablet OR computer OR app-based OR web-based OR handheld OR console OR device OR smartphone OR Kinect OR virtual reality OR controller OR augmented reality)) |
| 6 | "Games, Experimental"[Mesh] OR "Gamification"[Mesh] |
| 7 | game OR games OR gaming OR gamification |
| 9 | cogniti* OR memor* OR learn* OR executive function* OR language OR perceptual-motor function OR complex attention OR neurorehabilitation |
| 10 | "Cognition"[Mesh] OR "Brain"[Mesh] OR "Executive Function"[Mesh] OR "Psychomotor Performance"[Mesh] OR "Language"[Mesh] OR "Learning"[Mesh] OR "Attention"[Mesh] OR "Neurological Rehabilitation"[Mesh] |
| 12 | (#1 or #2) AND (#3 OR ((#4 OR #5) AND (#6 OR #7))) AND (#9 OR #10) |
| 13 | (#1 or #2) AND (#3 OR ((#4 OR #5) AND (#6 OR #7))) AND (#9 OR #10) |

**Appendix 2. Pubmed Search**
